# IgA dominates the early neutralizing antibody response to SARS-CoV-2

**DOI:** 10.1101/2020.06.10.20126532

**Authors:** Delphine Sterlin, Alexis Mathian, Makoto Miyara, Audrey Mohr, François Anna, Laetitia Claër, Paul Quentric, Jehane Fadlallah, Pascale Ghillani, Cary Gunn, Rick Hockett, Sasi Mudumba, Amélie Guihot, Charles-Edouard Luyt, Julien Mayaux, Alexandra Beurton, Salma Fourati, Jean-Marc Lacorte, Hans Yssel, Christophe Parizot, Karim Dorgham, Pierre Charneau, Zahir Amoura, Guy Gorochov

## Abstract

A major dogma in immunology has it that the IgM antibody response precedes secondary memory responses built on the production of IgG, IgA and, occasionaly, IgE. Here, we measured acute humoral responses to SARS-CoV-2, including the frequency of antibody-secreting cells and the presence of specific, neutralizing, antibodies in serum and broncho-alveolar fluid of 145 patients with COVID-19. Surprisingly, early SARS-CoV-2-specific humoral responses were found to be typically dominated by antibodies of the IgA isotype. Peripheral expansion of IgA-plasmablasts with mucosal-homing potential was detected shortly after the onset of symptoms and peaked during the third week of the disease. While the specific antibody response included IgG, IgM and IgA, the latter contributed to a much larger extent to virus neutralization, as compared to IgG. However, specific IgA serum levels notably decrease after one month of evolution. These results represent a challenging observation given the present uncertainty as to which kind of humoral response would optimally protect against re-infection, and whether vaccine regimens should consider boosting a potent, although, at least in blood, fading IgA response.

**One sentence Summary:** While early specific antibody response included IgG, IgM and IgA, the latter contributed to a much larger extent to virus neutralization.

## Introduction

In December 2019, a new coronavirus coined SARS-CoV-2 has emerged to cause an acute respiratory disease known as coronavirus disease 19 (COVID-19). The virus, identified as a betacoronavirus, spread worldwide with an unprecedented speed compared to the earlier Severe Acute Respiratory Syndrome Coronavirus (SARS-CoV) in 2003 and Middle East respiratory syndrome Coronavirus virus (MERS-CoV) in 2012 *(1)*. Recent reports indicate that SARS-CoV-2 elicites robust antibody responses, including specific IgG, IgA and IgM. Patients achieved a seroconversion within 20 days after symptoms onset, although with different kinetics of IgM and IgG production *(2–4)*.

Secretory IgA plays a crucial role in protecting mucosal surfaces against pathogens by neutralizing respiratory viruses or impeding their attachment to epithelial cells *(5–8)*. It has been demonstrated that influenza-specific IgA is more effective to prevent infections in mice and human than specific IgG. Elevated IgA serum levels have been correlated with influenza vaccine efficacy *(9–11)*. IgA might also play an important role in SARS-CoV infections. In mice, an intranasal delivery system of SARS-CoV proteins provide a better protection against SARS-CoV challenge than intramuscular administrations, suggesting that mucosal-induced IgA contributes efficiently against viral infection *(12)*. A recently reported intervention based on an intranasal immunization with a MERS-derived vaccine confirmed the beneficial role of IgA *(13)*. However, to which extent IgA production steps in to control natural SARS-CoV-2 infection in humans remains poorly understood.

Here, we tracked antibody-secreting cells in blood of SARS-CoV-2-infected patients. We then determined specific antibody titers in serum and studied their neutralizing capacities. Our results show that human IgA antibodies are often detectable before the appearance of SARS-CoV-2-specific IgG and argue in favor of a key role for IgA antibodies in early virus neutralization.

## Results

### Circulating plasmablasts preferentially express IgA1

The rapid, albeit transient, appearance of plasmablast in the blood circulation is a typical feature of the acute phase of viral infections *(14)*. We longitudinally monitored phenotypic changes of B cells in the blood of 38 SARS-CoV-2-infected patients (Table S1) using flow cytometry. Plasmablasts are immature antibody-secreting cells, defined here as cell cycling (Ki67^+^) CD19^low^CD27^high^CD38^high^ cells (Figure 1A). Their proportions very significantly increased among the B cell compartment in the early days of the first week after the onset of symptoms (median[min-max]%; 4.9[1.1-17.8]% *vs* 0.5[0.1-1.5]% in healthy donors; n=21 and n=9, respectively; Figure 1B), peak between days 10 to 15 (11.8[0.7-62.1]%, n=28, Figure 1B) and then decreased (4.4[0.2-33.8]% between day 16 to 25, n=21; 0.5[0.1-3.2]%, after day 50, n=14; Figure 1B). Longitudinal follow-up also confirmed the transient nature of this plasmablasts expansion in acute viral infection (Figure S1A).

**Figure 1:**
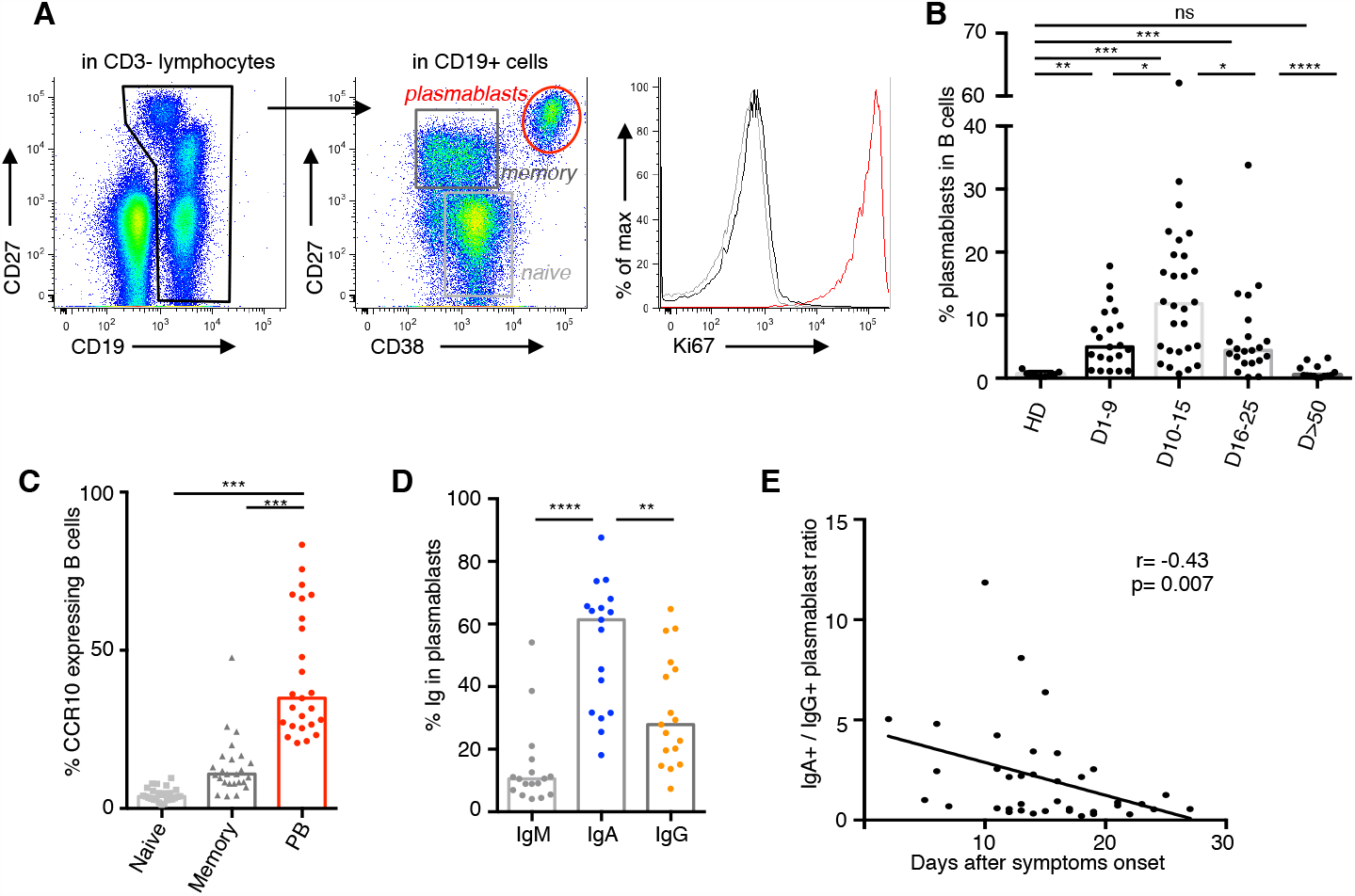
Plasmablasts dynamics following SARS-CoV-2 infection. A. Representative flow cytometry analysis of B-cell subpopulations in the blood of SARS-CoV-2 infected patients. Doublets and dead cells were excluded before CD3-CD19+ gating. Then, plasmablasts are defined as CD19^low^CD27^high^CD38^high^Ki67^+^ cells, memory B cells as CD19^+^CD27^+^IgD^-^Ki67^-^ and naïve B cells as CD19^+^CD27^-^ IgD^+^Ki67^-^ cells (IgD not shown). B. Plasmablasts frequency in B cells in blood of SARS-CoV-2 infected patients (n=38, clinical characteristics in Table S1) compared to healthy donors (n=9). Histograms represent medians. P values were calculated using Mann-Whitney test (* p<0.05; ** p<0.01; *** p<0.001). C. Flow cytometry analysis of CCR10 expression in B cell subpopulations in blood of SARS-CoV-2 infected patients (n=25). Samples used in this analysis were collected from day 3 to 27 after symptoms onset. Histograms represent medians. P values were calculated using Wilcoxon test (*** p<0.001). D. Intracellular antibody expression in circulating plasmablasts in blood of SARS-CoV-2 infected patients (n=17) using flow cytometry. Samples used in this analysis were collected from day 2 to 23 after symptoms onset. Histograms represent medians. P values were calculated using Wilcoxon test (** p<0.01; *** p<0.001). E. Intracellular IgA versus IgG expression in plasmablasts according to disease duration. Each dot represents one patient. Non-parametric Spearman correlation was calculated.

We probed these circulating plasmablasts for their surface expression of CCR10, a chemokine receptor involved in the migration of various immune cells to mucosal sites, especially the lung *(15, 16)*. Less than 10% of memory and naive B cells, but approximately 40% of detected plasmablasts, were CCR10+ (3.8[1.2-9.6]% in naive B cells *vs* 10.9[4.1-25.9]% in memory B cells *vs* 34.9[20.7-83.4]% in plasmablasts; n=25; Figure 1C), suggesting a substantial lung tissue tropism of the latter. Although we analyzed the early phase of the immune response, only a minor population of plasmablasts was found to be IgM producers, as measured using intracellular staining (10.5[4.2-54.1]% IgM^+^ plasmablasts; n=17; Figure 1D). In contrast, most plasmablasts expressed IgA (61.4[18.1-87.6]% IgA^+^ plasmablasts *vs* 27.9[7.4-64.8]% IgG^+^ plasmablasts; n=17; Figure 1D), a feature consistent with a mucosal phenotype for these cells. Intracellular IgA subclass identification showed higher frequencies of IgA1-expressing plasmablasts, as compared to IgA2 (66[26.8-88.5]% IgA1^+^ vs 31.6[3.7-70.8]% IgA2^+^ in IgA^+^ plasmablasts; n=13; Figure S1B-C). This first wave of circulating IgA-expressing plasmablasts was followed by a second wave of IgG-expressing cells that became predominant by day 22 after the onset of symptoms (Figure 1E, S1D-E). While the majority of IgA^+^ expressed CCR10, this marker was expressed by only a minority of IgG^+^ plasmablasts (60.5[37.6-92.6]% *vs* 23.3[3.2-78]% CCR10+, n=15; Figure S1F), suggesting that the latter may niche differently, and most likely in the bone marrow. Of note, the frequency of peripheral IgM-expressing plasmablasts did not significantly vary with time (Figure S1G) and only marginally at later time points (Figure S1H).

In a recent study that characterized the immune response of a COVID-19 patient, the induction of T follicular helper (Tfh) cells was reported to occur concomitantly with that of plasmablasts *(17)*. In order to evaluate a potential germinal center origin of the plasmablast wave observed in our patients, we longitudinally tracked CD4^+^CXCR5^+^PD1^+/-^ Tfh cells in their blood. We found no significant increase in the frequency of Tfh subsets in COVID-19 patients, as compared to healthy donors, at any of the analyzed time points (Figure S2A-B). Neither activated (CD4^+^CXCR5^+^PD1^+^), nor latent (CD4^+^CXCR5^+^PD1^-^), Tfh cells were correlated with plasmablasts frequency (Figure S2C). The frequency of neither activated (CD4^+^CXCR5^+^PD1^+^), nor latent (CD4^+^CXCR5^+^PD1^-^) Tfh cells was found to correlate with that of plasmablasts. (Figure S2C).

Taken together, these results point toward an early humoral response to SARS-CoV-2 dominated by IgA-expressing plasmablasts, circulating across mucosal sites.

### Early SARS-CoV-2-specific IgA detection

Photonic ring immunoassay is a novel technology that allows to simultaneously test in parallel various antigens (Figure S3)*(18, 19)*. Using this technique, we assessed the prevalence of IgG, IgA and IgM antibodies recognizing the SARS-CoV-2 full-length nucleocapsid protein (NC) or Spike Receptor-binding domain (S1-RBD) in 214 serum samples of 135 infected patients (Tables S2 and S3).

The appearance of IgA or IgG antibodies directed at SARS-CoV-2 S1-RBD was detected, in only three cases, as early as day 1 and 2 after the onset of the first symptoms, respectively (Figure S4). Anti-S1-RBD IgG and IgA were detected in 7 and 15 out of 48 samples, respectively, during the first week (Figure 2A and Figure S4). Although IgM is typically considered as a marker of acute infection, anti-S1-RBD IgM were detected only in 7 out of these 48 early samples (Figure 2A and Figure S4). Moreover, anti-NC IgM remained undetectable in all samples except one.

**Figure 2:**
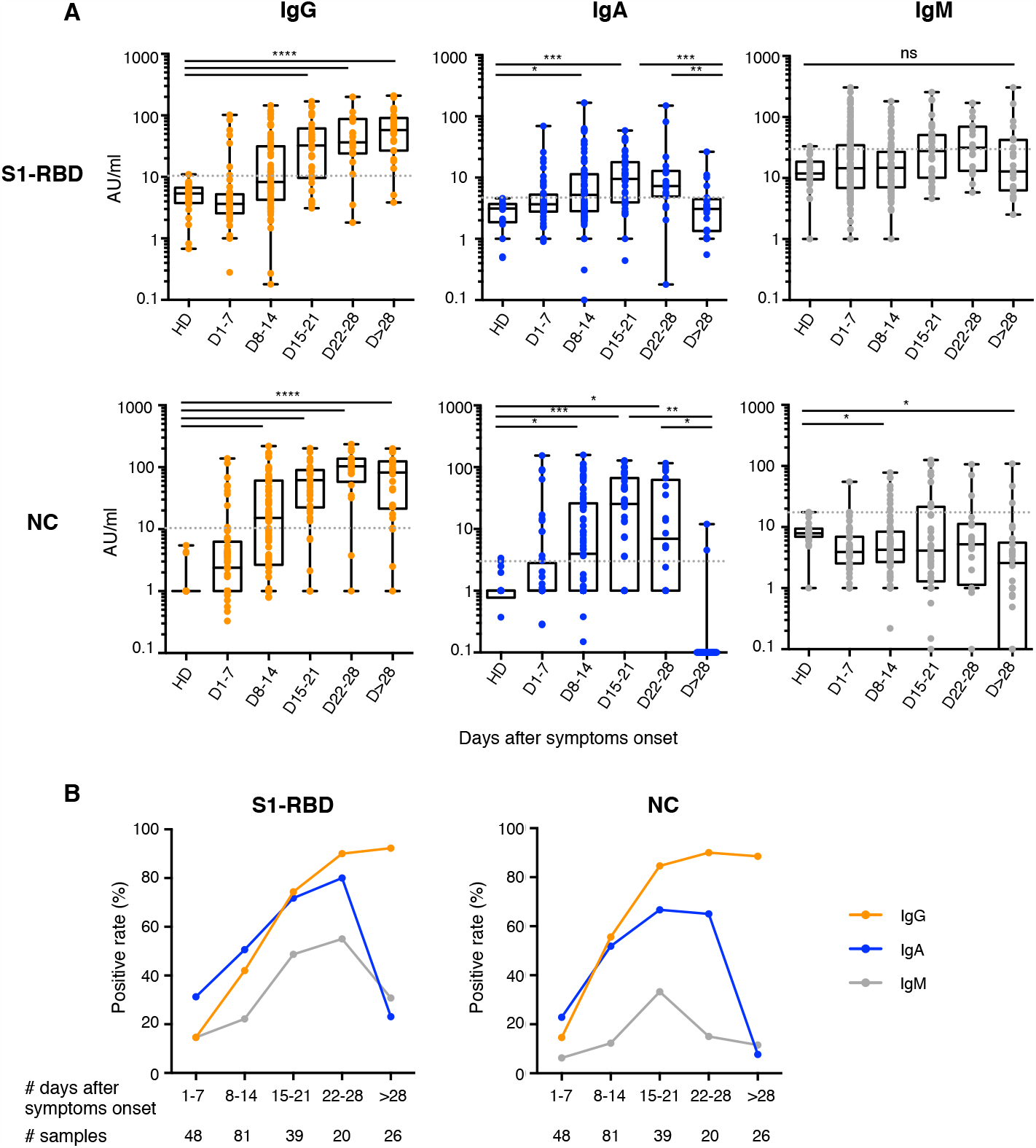
Antibody responses kinetics to SARS-CoV-2 proteins. A. Specific IgG, IgA and IgM against Spike-1 Receptor Binding Domain (S1-RBD) and Nucleocapsid protein (NC) were measured using photonic ring immunoassay in 214 sera from 135 patients (clinical characteristics detailed in Tables S2-S3). Antibody levels are expressed as arbitrary units/ml (AU/ml). Cut-off lines are represented as grey dotted lines. The boxplots show medians (middle line) and first and third quartiles while the whiskers indicate minimal and maximal values. P value was calculated using Kruskal-Wallis test (* p<0.05; **p<0.01: ***p<0.001; ****p<0.0001). B. Positive rates of specific IgG, IgA and IgM in 214 sera at different times after symptoms onset, from day 1 to 78.

The proportion of patients with detectable anti-S1-RBD IgG increased until reaching a plateau approximately during the fourth week after symptoms onset (positive samples: 15% day 1-7; 42% day 8-14; 74% day 15-21; 90% day 22-28 and 92% day>28; Figure 2A-B). In contrast, the frequency of patients with anti-S1-RBD IgA peaked around day 22 (positive samples: 31% day 1-7; 51% day 8-14; 72% day 15-21 and 80% day 22-28; Figure 2A-B), then decreased by day 25 (positive samples: 23% day>28; Figure 2A-B). Following a similar kinetics with respect to the appearance of anti-S1-RBD antibodies, the proportion of patients with detectable anti-NC IgG remained stable around the fourth week post-symptoms onset (positive samples: 15% day 1-7; 56% day 8-14; 85% day 15-21; 90% day 22-28 and 89% day>28; Figure 2A-B) whereas anti-NC IgA quickly disappeared and were no longer detectable in most patients one month after disease onset (positive samples: 23% day 1-7; 52% day 8-14; 67% day 15-21; 65% day 22-28 and 8% day>28; Figure 2A-B). Of note, in two patients who recovered from COVID-19, no specific antibodies were detected at the available late time-points (Figure 2A).

Both anti-S1-RBD and anti-NC IgG titers increased over time (anti-S1-RBD arbitrary units: 3.6[0-102.1] day 1-7; 8.3[0.2-145.2] day 8-14; 32.6[3.5-168.9] day 15-21; 36.5[1.8-200.9] day 22-28; 57.9[5.1-209.9] day>28; anti-NC arbitrary units: 2.4[0-138.2] day 1-7; 15.2[0-219.8] day 8-14; 61.7[0-201.7] day 15-21; 103.5[0-236.2] day 22-28; 82.4[0-200.2] day>28; Figure 2A) while virus-specific IgA titers rised during the first 3 weeks after symptoms onset, then dropped and were undetectable one month after recovery (anti-S1-RBD arbitrary units: 3.7[0-69.6] day 1-7; 5.2[0-166.3] day 8-14; 9.5[0-58.5] day 15-21; 7.3[0.2-149.9] day 22-28; 3.1[0-26.4] day>28; anti-NC arbitrary units: 0[0-153.7] day 1-7; 3.9[0-158.2] day 8-14; 25.5[0-128.2] day 15-21; 6.9[0-116.6] day 22-28; 0[0-11.9] day>28; Figure 2A).

Altogether, these results suggest that anti-SARS-CoV-2 IgA testing should improve early COVID19 diagnosis, while testing serum more than 28 days after the onset of symptoms should mainly rely on the detetion of IgG antibodies.

### IgA is a potent SARS-CoV-2 neutralizing agent

We then sought to determine the respective contribution of each of the IgG and IgA isotypes to virus neutralization. We assessed the neutralizing capacity of serum antibodies using a pseudoneutralization assay. The neutralization potential of serum, tested at dilution 1/40 according to previous studies rapidly increased during disease course, and plateaued by day 10 post-symptoms onset (Figure 3A and Figure S5A). Neutralizing activity, determined in 12 sera having reached this plateau, was found to vary considerably between patients with half maximal inhibitory concentration (IC_50_) values ranging from 1/169 to 1/16189 serum dilution (Figure S5B).

**Figure 3:**
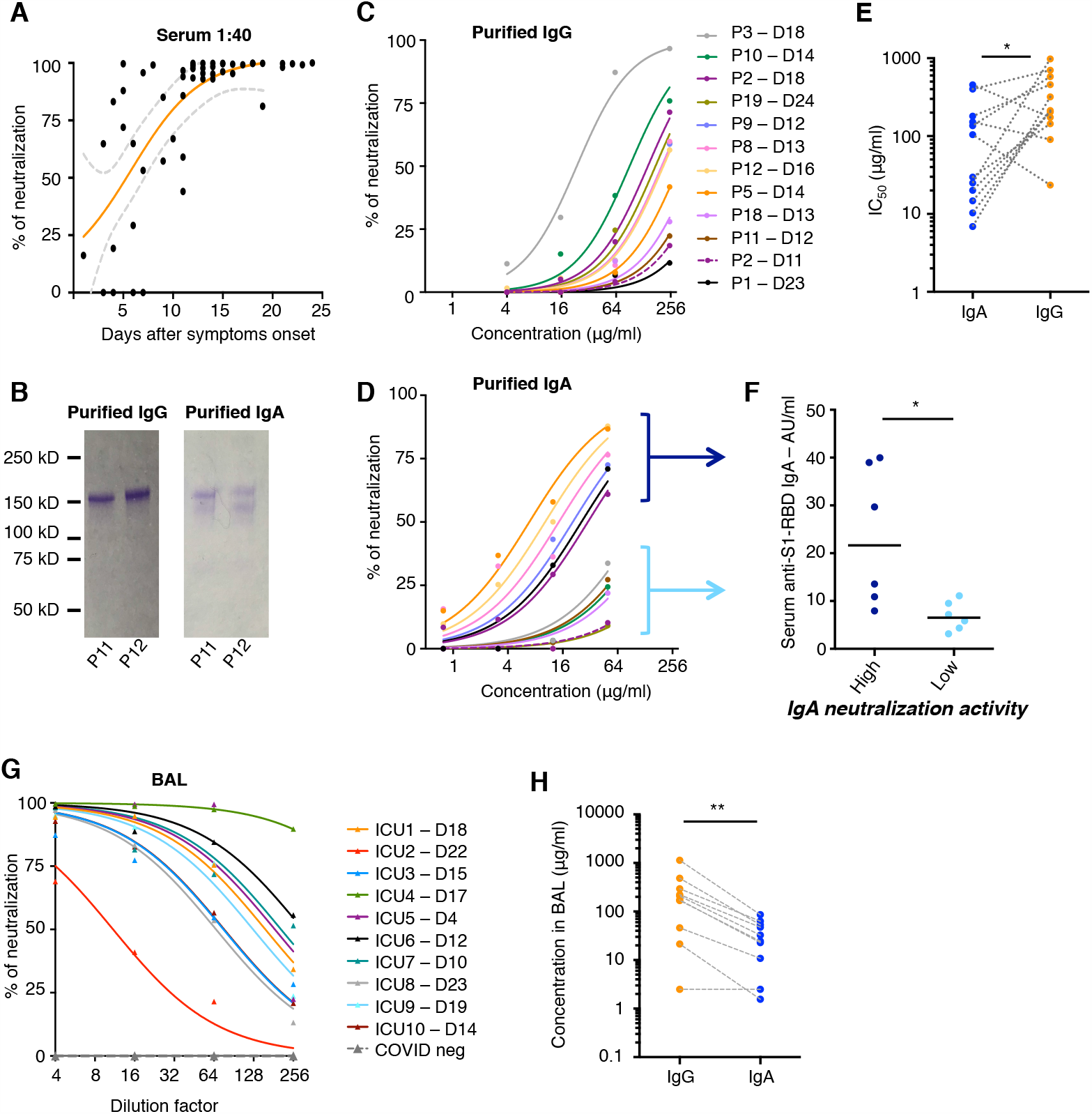
Neutralizing activity of serum antibodies to SARS-CoV-2. A. Neutralizing activity of 52 sera (dilution 1:40) from 38 SARS-CoV-2 infected patients (see clinical characteristics in Table S1) was determined by pseudovirus neutralisation assay. Orange curve represents significant sigmoidal interpolation (p=0.0082). Grey dotted lines represent 95% confidence intervals curves. B. Purified IgG and IgA are monomeric. Representative SDS-PAGE showing purified IgG and IgA in non-reducing conditions for P11 and P12. The same experiment was repeated for the 12 purified IgG and IgA pairs. C. Neutralizing activity of purified IgG was measured at indicated concentrations from 12 sera collected between day 11 and day 24 post-symptoms onset (as indicated). Curves were drawn according to non-linear regression. D. Neutralizing activity of purified IgA from paired Fig. 3C samples, analysed as in Fig. 3C. High and low IgA neutralization potential are indicated with dark and light blue brackets. E. Compaired purified IgA and IgG IC_50_ values in samples tested in Figs. 3C and D. P value was calculated using Wilcoxon test (* p<0.05). F. Comparison of anti-S1-RBD IgA levels measured by photonic ring immunoassay in serum showing high (IC_50_<30μg/ml) versus low IgA (IC_50_>100μg/ml) neutralizing activity. P value was calculated using Mann-Whitney test (* p<0.05). G. Neutralizing activity of bronchoalveolar lavages (BAL) collected in 10 SARS-CoV-2 patients between day 4 and 23 after symptoms onset (clinical characteristics are detailed in Table S4). Indicated BAL dilutions were tested using pseudovirus neutralization assay. Bronchoalveolar lavages obtained from SARS-CoV-2 negative patients (n=3) showed no neutralization activity (dotted grey lines). Each colored line represent one patient. H. IgG and IgA levels measured by ELISA in bronchoalveolar lavages tested in panel F. P value was calculated using Mann-Whitney test (** p<0.01).

To detail the respective contributions of the dominant antibody isotypes to virus neutralization, purified IgA and IgG fractions from the serum of 12 patients (Figure 3B) were back-to-back tested for their neutralizing capacity (Figure 3C, D). Strikingly, IgA were far more potent in their capacity to neutralize SARS-CoV-2, as compared to paired IgG (Figure 3C-E). While IgG were able to neutralize the virus only at undiluted serum concentrations, purified IgA fractions had approximately five times lower IC_50_s, as compared to purified IgG ([min-max]; IgA IC_50_ [6.9-454.9] vs IgG IC_50_ [23.6-982.4]; n=12, Figure 3E). In our cohort, two profiles of sera could be distinguished based on their high or low IgA neutralization potential (Figure 3D). Interestingly, these profiles were directly associated with lower or higher anti-S1-RBD IgA serum titers, respectively (6.5[3.1-11.1] *vs* 21.6[7.9-38.9], n=12, p=0.02; Figure 3F), suggesting that, as expected, IgA neutralisation potential is mainly relying on RBD targeting. Importantly, the more efficient neutralisation potential of IgA compared to IgG cannot be explained by an avidity effect, as both purified antibody preparations were monomeric (Figure 3B).

These observations highlight the neutralization potential of systemic humoral immunity driven by both IgA and IgG. However, the main SARS-CoV-2 targets are lung epithelial cells *(20, 21)*, and mucosal immunity differs from systemic immunity. Therefore, to assess local immunity, bronchoalveolar lavage (BAL) samples obtained from 10 patients (Table S4) were tested for their neutralization potential (Figure 3G). As shown, BAL samples harvested at various times, and as early as 4 days after symptoms onset, harbored detectable SARS-CoV-2 neutralization activity, suggesting the mounting of an efficient local immune response as well. Of note, IgG concentrations were always superior than those of IgA in the tested BAL samples, except one (Figure 3H).

## Discussion

We have studied the antibody response of COVID-19 patients and show that SARS-CoV-2 infection induces an early and potent virus-specific IgA response that precedes the production of SARS-CoV-2 IgG. Importantly, IgA antibodies purified from the serum of these patients were found to more efficiently neutralize SARS-CoV-2 *in vitro* than IgG. However, we also observed a rapid decline in SARS-CoV-2-specific IgA serum levels, thereby questioning the long-term efficacy of this first wave response, as efficient as it appears to be. It remains nevertheless possible that sustained SARS-CoV-2-specific secretory IgA levels are maintained in secretions, and particularly in the pulmonary lumen. Indeed, previously documented re-circulating IgA-secreting plasmablasts with a mucosal-homing profile were detected in high numbers in the patients we studied, and could seed their lung/airway interface *(16, 22–24)*. It is also known that IgA-secreting cells efficiently home to and reside within human mucosa *(25)*, and that IgA subclass switch recombination can directly take place in this tissue *(26)* in a T cell-independent manner *(27)*. The lack of correlation between plasmablast and Tfh cell expansion, observed in the present study, could argue in favor of a prominent germinal center-independent induction of IgA *(28)*. Of note, several recently described SARS-CoV-2 neutralizing IgG *(29, 30)* were devoid of somatic mutations typically associated with affinity maturation and T cell help. In this respect, a molecular and functional characterisation of IgA monoclonal antibodies secreted by plasmablasts accessible in blood during the first week of symptoms should shed light on their mutational status. The observation made in our study could also give clues to explain the puzzling observation that the vast majority of children are not afffected by COVID-19 *(31, 32)*, by postulating that cross-reactive IgA, recently identified in human gut mucosa against other targets than SARS-CoV-2 *(22, 33)*, might be more prevalent in children and/or could be rapidly mobilized in response to infection with SARS-CoV-2.

The BAL samples tested in this study were obviously obtained from severe COVID-19 patients, all affected with severe lung damage, and therefore possible serum contamination. It remains to be confirmed whether the BAL IgG prevalence observed (Figure 3H), is representative of the pulmonary humoral status of mild and convalescent patients. To address this issue, it is important to evaluate, in future studies, the presence of SARS-CoV-2-specific secretory IgA at easily accessible sites, such as in saliva from COVID-19-recovered patients. Saliva probing might become technically feasible given the exquisite sensitivity of novel digital ELISA-based assays for SARS-CoV-2-specific IgA detection *(34)*. Future studies will also confirm the suitability of the photonic ring immunoassay technology, used in the present study, for the detection of IgA in saliva. The latter approach could represent an attractive alternative for the mass testing programs that need to be implemented, given that results are rapidly obtained using a small-size fully automated instrument, and only a few microliters of sample volume.

It is also important to note that in some of the early serum samples with efficient virus neutralizing capacity, only anti-RBD IgM, but neither IgA nor IgG SARS-CoV-2 spike RBD-specific antibodies were above the detection threshold (P2 day 14 and P3 day 6 post symptoms, Figure S5C). This observation points to a protective potential for IgM as well. The latter sequence (M first, then G and A) is however not likely to be prevalent. A more typical profile is exemplified by patient P9, with anti-RBD IgA levels peaking before the appearance of anti-RBD IgG, and barely detectable IgM at any time point (Figure S5C). Since, in rare cases, only specific IgM are detected at early time points, it remains recommended to measure the presence of all isotypes for serological diagnosis.

Our study presents several limitations. Functional mucosal immunity analysis was only carried out in BAL, and longitudinal studies are needed at various body sites in order to assess whether local SARS-CoV-2-specific IgA production might be more persistant than in blood. In addition, it remains to be confirmed whether the *in vitro* IgA neutralization efficacy of the purified IgA serum antibodies translates into a potent barrier effect, not only in recovered patients, but also in paucisymptomatic individuals and healthy carriers.

In conclusion, we would like to stress the importance of mucosal immunity as an important defence mechanism against SARS-CoV-2 to be monitored in infected patients, as well as to recommend testing the usefulness of a vaccine protocol aimed at inducing a specific respiratory IgA response to SARS-CoV-2.

## Materials and methods

### Patients recruitment and samples preparation

Fresh blood sample from 135 consecutive adult patients with COVID-19 referred to the Department of Internal Medicine 2, Pitié-Salpêtrière Hospital, Paris were included in the study between March 22, 2020 and April 24, 2020 and compared with 20 age and sex-matched healthy donors (HDs). The diagnosis of COVID-19 relied on SARS-CoV-2 carriage in the nasopharyngeal swab, as confirmed by real-time reverse transcription-PCR analysis. Ten additional patients with chest computed tomography (CT) scan displaying features suggesting a COVID-19 infection and tested positive for the presence of serum anti-SARS-CoV-2 antibodies were also included. Demographic and clinical characteristics are detailed in Tables S1-S3. Broncho-alveolar lavages were collected from 10 COVID-19 patients hospitalized in Intensive Care Units, Pitié-Salpêtrière Hospital, Paris and compared with 3 COVID-19-negative samples. Demographic and clinical characteristics are detailed in Tables S4. All patients gave informed consent. This study was approved by the local ethical committee of Sorbonne Université (n°2020-CER2020-21). For all patients, sera were stored immediately after collection at −80°C. Peripheral blood mononuclear cells (PBMCs) were isolated from blood samples of 38 patients after Ficoll-Hypaque density gradient centrifugation (Eurobio, Courtaboeuf, France) and analyzed immediately. Clinical characteristics of these patients are presented in Table S1.

### B-cell and T-cell phenotyping

Phenotyping was assessed on freshly isolated PBMCs stained with a combination of anti-human antibodies (Table S5). Intracellular staining was performed on fixed and permeabilized cells (using the FOXP3 Transcription Factor Staining Buffer kit, eBioscience). Cells were acquired on a BD FACSCanto II Flow cytometer (BD biosciences) and analysed with FlowJo v10 software (FlowJo, LLC).

### Serological analysis

SARS-CoV-2 specific IgA, IgM and IgG antibodies were measured in 214 serum samples from 135 patients with The Maverick (tm) SARS-CoV-2 Multi-Antigen Serology Panel (Genalyte Inc. USA) according to the manufacturer’s instructions. The Maverick (tm) SARS-CoV-2 Multi-Antigen Serology Panel (Genalyte Inc) is designed to detect antibodies to five SARS-CoV-2 antigens: nucleocapsid, Spike S1 RBD, Spike S1S2, Spike S2 and Spike S1 with in a multiplex format based on photonic ring resonance technology *(18, 19, 35)*. This system detects and measure with good reproducibility (Figure S3) changes in resonance when antibodies bind to their respective antigens in the chip. The instrument automates the assay. Briefly, 10µl of each serum samples were added in a sample well plate array containing required diluents and buffers. The plate and chip are loaded in the instrument. First the chip is equilibrated with the diluent buffer to get baseline resonance. Serum sample is then charged over the chip to bind specific antibodies to antigens present on the chip. Next, chip is washed to remove low affinity binders. Finally, specific antibodies of patients are detected with anti-IgG or -IgA or -IgM secondary antibodies. 20 sera collected before December 2019 were analyzed to calculate cut-off values. Positivity was defined as results above the 99^th^ percentile.

### Purification and quantification of IgA and IgG from serum

IgA and IgG were isolated from 12 serum samples diluted in 1X-PBS as previously described *(33)*. Sera were selected after SARS-CoV-2 specific antibodies evaluation. Briefly, serum samples were load onto peptide M/Agarose or Protein G/Agarose column (Invivogen) after column equilibration. Chromatography steps were performed at a flow rate of 0.5ml/min. Next, 20 column volumes of 1X-PBS was used to wash the column. IgA and IgG were then eluted with 5ml of 0.1M glycine (Sigma-Aldrich) and pH was immediately adjusted to 7.5 with 1M Tris. 1X-PBS buffer exchange was achieved using Amicon® Ultra centrifugal filters (Merck Millipore) through a 100-kD membrane according to the manufacturer’s instructions. The quantification of IgA and IgG was determining using NanoVue Plus microvolume spectrophotometers.

### Pseudovirus production and permissive cell line generation

Pseudotyped vectors were produced by triple transfection of 293T cells as previously described *(36)*. Briefly, cells were co-transfected with plasmids encoding for lentiviral proteins, a luciferase Firefly reporter and plasmid expressing a codon-optimized SARS-CoV-2 Spike gene. Pseudotyped vector were harvested at day 2 post-transfection. Functional titer (TU) was determined by qPCR after transduction of a stable HEK 293T-hACE2 cell line. To generate this cell line, HEK 293T cells were transduced at MOI 20 with an integrative lentiviral vector expressing human ACE2 gene under the control of UBC promoter. Clones were generated by limiting dilution and selected on their permissivity to SARS-CoV-2 S pseudotyped lentiviral vector transduction.

### Pseudoneutralization Assay

First, serum dilutions are mixed and co-incubated with 300 TU of pseudotyped vector at room temperature during 30 minutes. Serum and vector are diluted in culture medium (DMEM-glutamax (Gibco) + 10% FCS (Gibco) + Pen/Strep (Gibco). Mix is then plated in Tissue Culture treated black 96-well plate (Costar) with 20 000 HEK 293T-hACE2 cells in suspension. To prepare the suspension, cell flask is washed with DPBS twice (Gibco) and cell are individualized with DPBS + 0.1% EDTA (Promega) to preserve hACE2 protein. After 48h, medium is removed and bioluminescence is measured using a Luciferase Assay System (Promega) on an EnSpire plate reader (PerkinElmer).

### Statistical analysis

Variables are expressed as the median. Nonparametric test were used as Mann-Whitney U test to compare two independent groups, Wilcoxon test to compare paired values and Spearman correlation test. Significant P values are indicated as described below: * p < 0.05; **p < 0.01; ***p < 0.001; ****p < 0.0001. Statistical analysis was performed using GraphPad Prism software, V6 (GraphPad, San Diego).

## Data Availability

All data related to this study are present in the paper or Supplementary Materials.

## Supplementary Materials

Figure S1 (related to Figure 1): Intracellular antibody expression in circulating plasmablasts.

Figure S2 (related to Figure 1): Circulating follicular helper T cells in blood of SARS-CoV-2 patients.

Figure S3 (related to Figure 2): Performance of photonic ring immunoassay to detect anti-S1-RBD and anti-NC antibodies

Figure S4 (related to Figure 2): Early detection of anti-S1-RBD antibodies in serum from SARS-CoV-2 patients.

Figure S5 (related to Figure 3): Neutralizing activity of serum from SARS-CoV-2 patients.

Table S1: Demographics, baseline characteristics, treatment and outcome of 38 patients with COVID-19 assessed for blood plasmablasts.

Table S2: Demographics and baseline characteristics of patients with COVID-19.

Table S3: Clinical characteristics, laboratory results, treatment and outcome of patients with COVID-19.

Table S4: Demographics, baseline characteristics, treatment and outcome of patients with acute respiratory distress syndrome during the course of COVID-19.

Table S5: Human Antibodies used for B and T cell phenotyping.

## Acknowledgments

The authors wish to thank the patients that agreed to participate in this study, doctors and nurses from the endocrinology, metabolism, nutrition and diabetology Departments of Institut E3M (Assistance Publique Hôpitaux de Paris (AP-HP), Hôpital Pitié-Salpêtrière, 75013 Paris, France) who participated in this study, all members from the Immunology Department, (Assistance Publique Hôpitaux de Paris (AP-HP), Hôpital Pitié-Salpêtrière, 75013 Paris, France), who volunteered to biobank blood and BAL samples, administrative staff at the Research and Innovation office of Pitié-Salpêtrière Hospital for their support, Laura Wakselman from clinical research unit (URC) of Pitié-Salpêtrière Hospital for help with regulatory and ethical issues, and A. Neumann for discussions and ideas.

## Author contributions

A.Mathian, J.F., A.G. C.E.L., J.M., A.B., Z.A., recruited patients. A.Mathian, J.F., P.Q., A.G. C.E.L., J.M., Z.A., collected demographic and clinical data. D.S., A.Mohr, L.C., P.Q., C.P., K.D., J.M.L., P.G., prepared the specimens. D.S., M.M., A.Mohr, L.C., K.D. designed and performed experiments. F.A. and P.C. designed, performed and analyzed neutralization assays. D.S., A.Mohr, L.C., F.A., analyzed data. D.S., A.Mohr, F.A. prepared the figures. D.S. and G.G. wrote the manuscript draft. D.S., A.Mathian, M.M., A. Mohr, J.M.L., Z.A., G.G. designed the study and reviewed the manuscript.

## Supplementary Materials

**Figure S1 (related to Figure 1):**
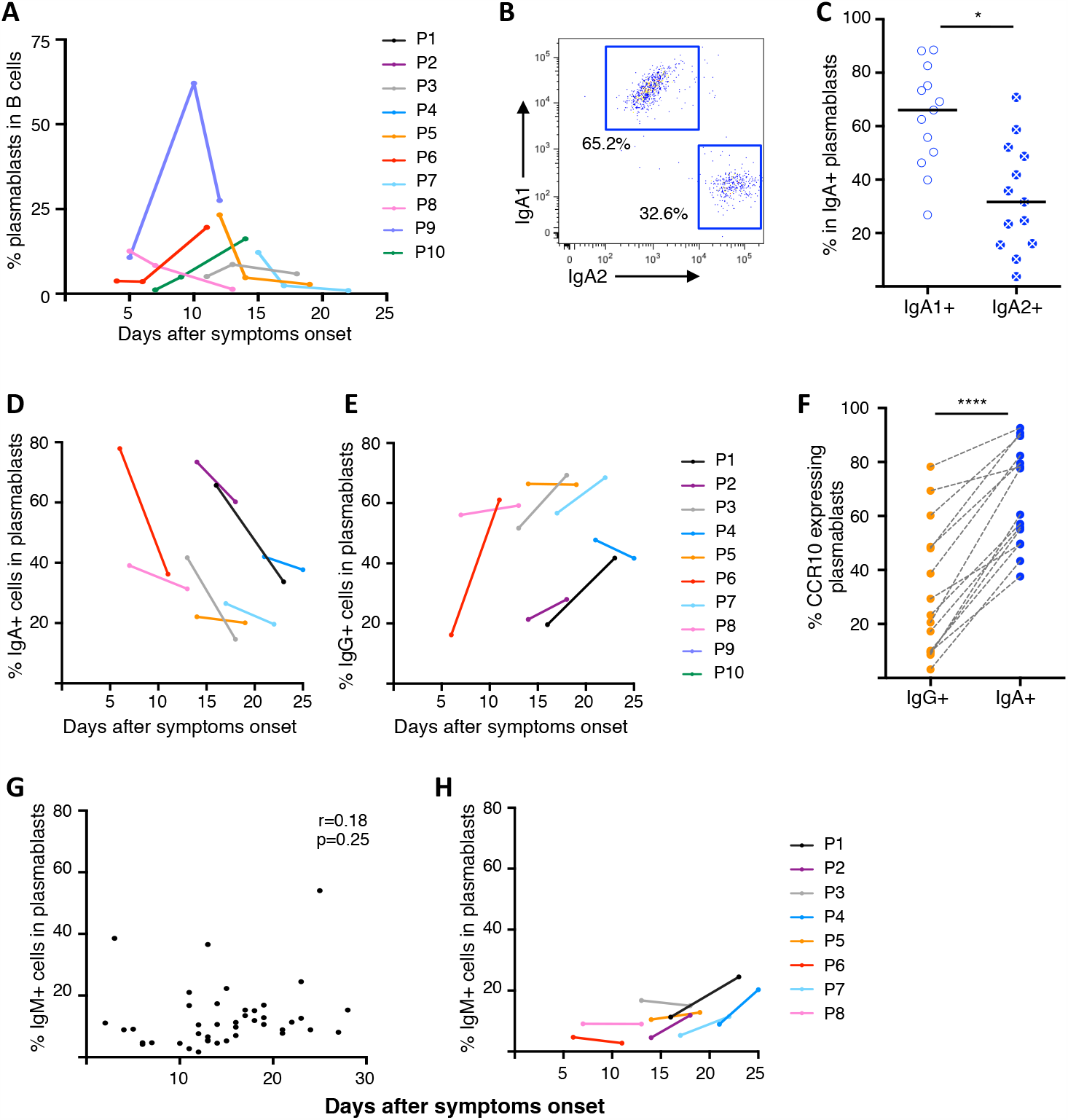
Intracellular antibody expression in circulating plasmablasts. A. Frequency of CD19^low^CD27^high^CD38^high^Ki67^+^ plasmablasts in B cells (gated on CD3-lymphocytes) measured by flow cytometry in blood collected at different time points after symptoms onset in 7 SARS-CoV-2 infected patients. Each colored line represents one patient. B. Intracellular IgA expression in plasmablasts measured by flow cytometry in blood collected at two different time points after symptoms onset in 8 SARS-CoV-2 infected patients. Each colored line represents one patient. C. Intracellular IgG expression in plasmablasts measured by flow cytometry in blood collected at two different time points after symptoms onset in 8 SARS-CoV-2 infected patients. Each colored line represents one patient. D. One representative flow cytometry analysis out of 13 of intracellular IgA1 and IgA2 expression in circulating IgA^+^ plasmablasts from one SARS-CoV-2 infected patient. E. Intracellular IgA subclass expression in IgA^+^ plasmablasts measured by flow cytometry in blood collected between day 2 and 23 after symptoms onset (n=13). F. Flow cytometry analysis of CCR10 expression in IgG^+^ and IgA^+^ plasmablasts in blood of SARS-CoV-2 infected patients (n=15). Samples used in this analysis were collected between day 3 and 27 after symptoms onset. P values were calculated using Wilcoxon test (*** p<0.001). G. Frequency of IgM-expressing cells among plasmablasts following SARS-CoV-2 disease onset. Each dot represents one sample (n=42). Non-parametric Spearman correlation was calculated. H. Longitudinal tracking of IgM-expressing cells among plasmablasts measured after intra-cellular staining and flow cytometry analysis in blood collected at indicated time points after symptoms onset in 8 SARS-CoV-2 infected patients. Each colored line represents one patient.

**Figure S2 (related to Figure 1):**
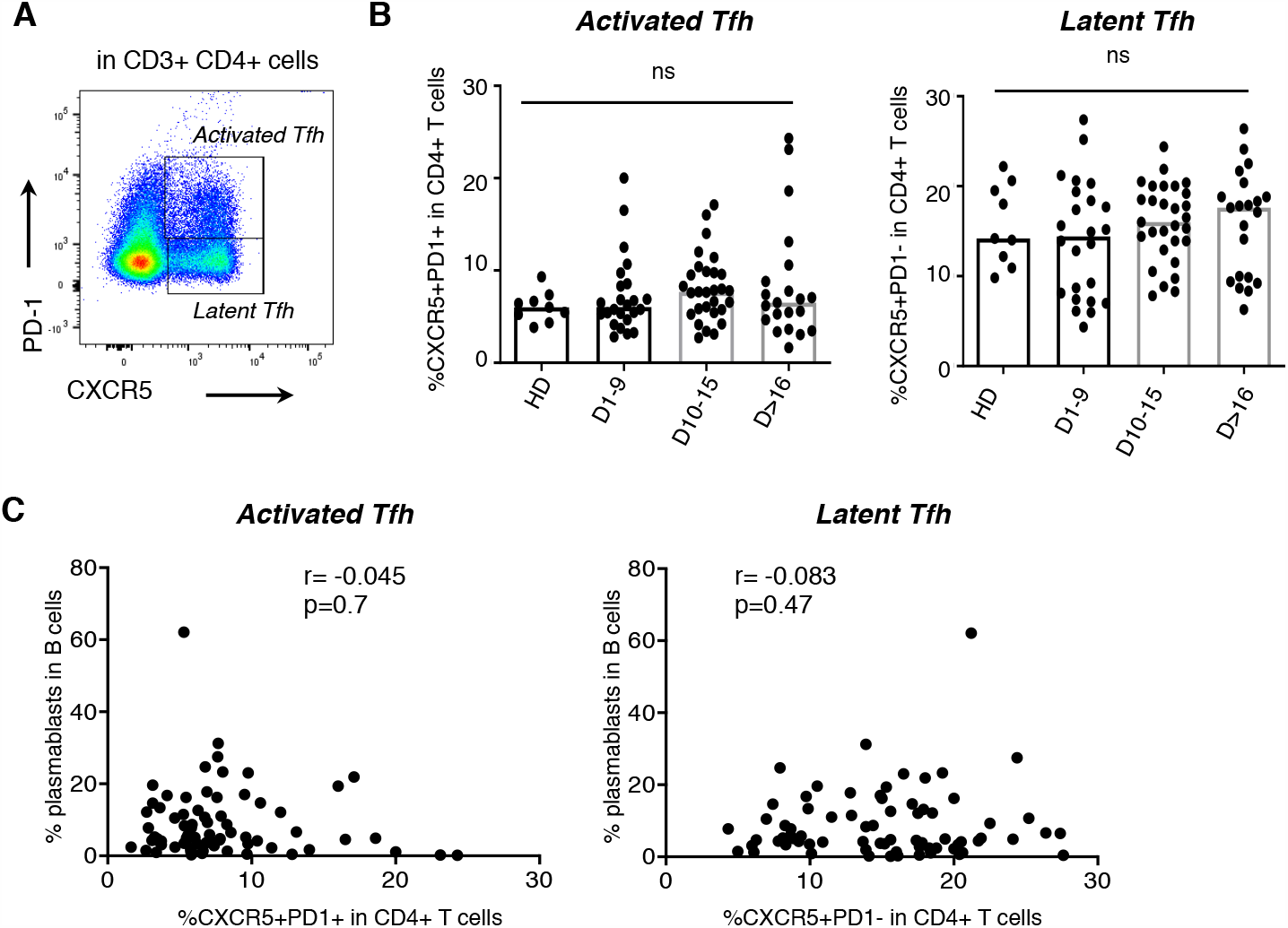
Circulating follicular helper T cells in blood of SARS-CoV-2 patients. A. Representative flow cytometry analysis of activated (CXCR5^+^PD-1^+^) and latent (CXCR5^+^PD-1^-^) follicular helper T cells, gated on CD3^+^CD4^+^ cells. B. Flow cytometry analysis of activated (left) and latent (right) Tfh cells in CD4^+^ T cells in blood of SARS-CoV-2 infected patients. Histograms represent medians. No significant (ns) difference was observed using Mann-Whitney test. C. Frequencies of activated (left) and latent (right) Tfh cells in CD4^+^ T cells were compared to the frequency of plasmablasts in B cells. Spearman coefficient (r) and p value (p) are indicated.

**Figure S3 (related to Figure 2):**
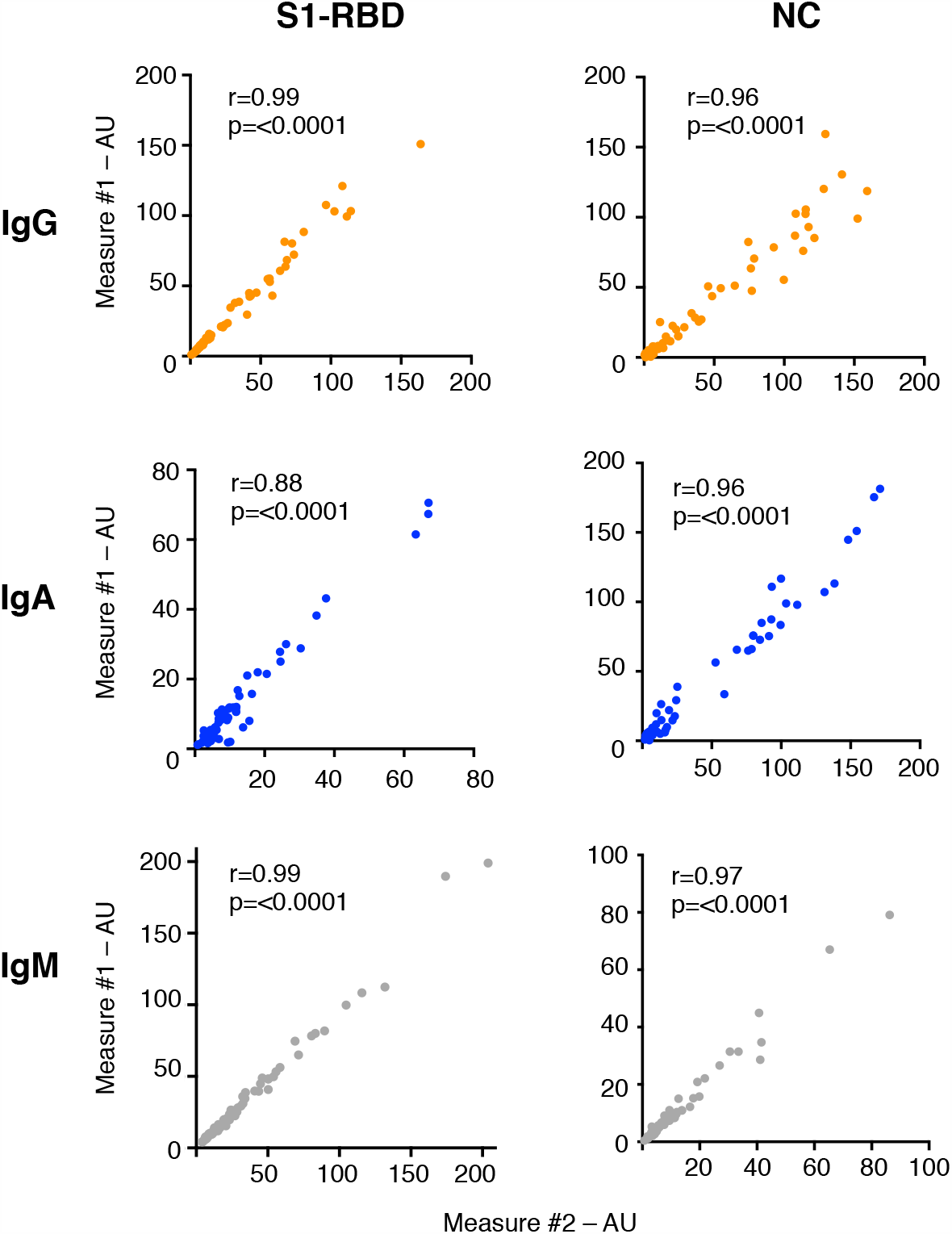
Performance of photonic ring immunoassay to detect anti-S1-RBD and anti-NC antibodies. Correlation analysis of IgG, IgA and IgM titers from 2 different measures in 74 sera from SARS-CoV-2 patients. Spearman coefficient (r) and p value (p) are indicated in graphs.

**Figure S4 (related to Figure 2):**
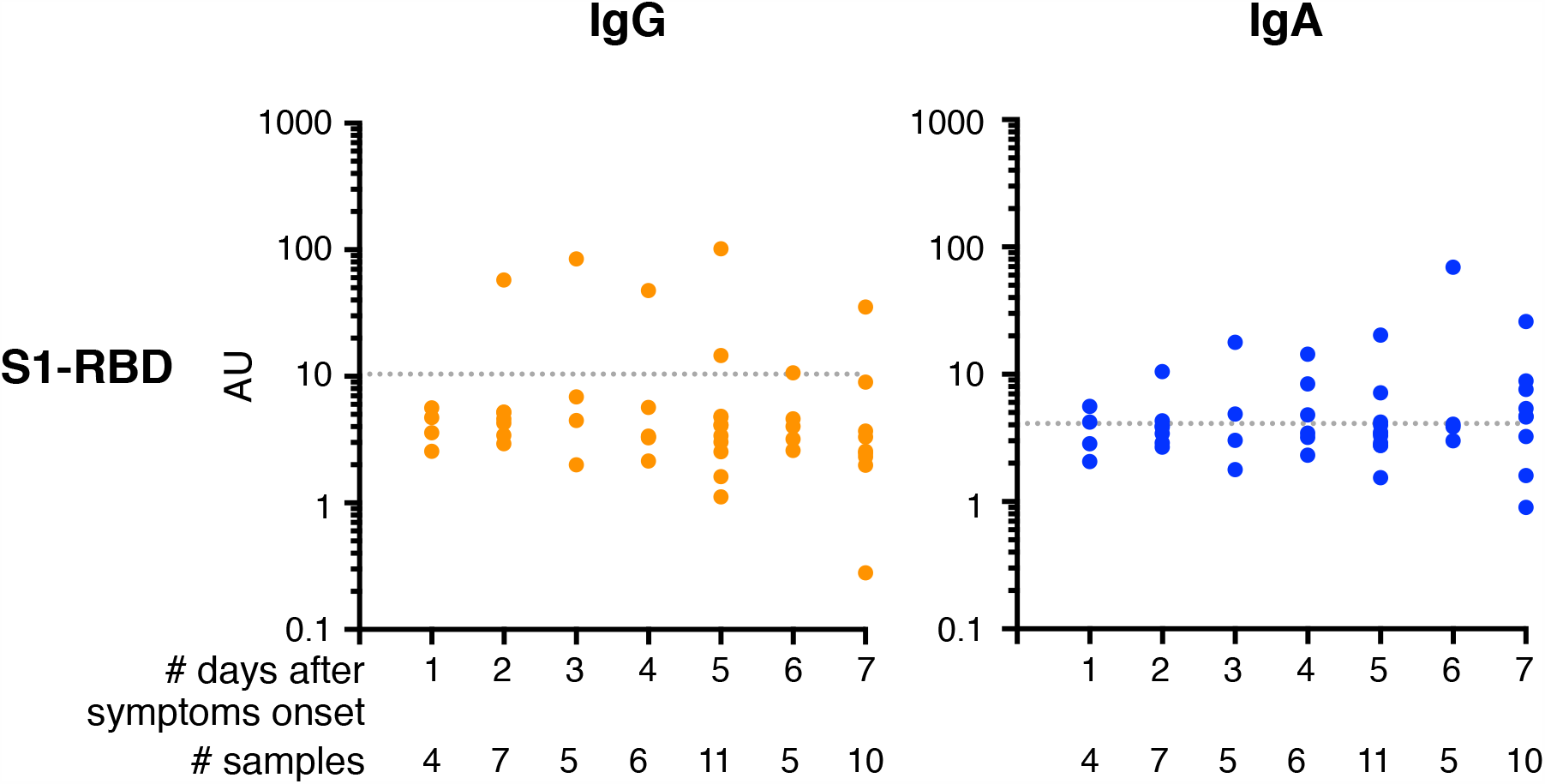
Early detection of anti-S1-RBD antibodies in serum from SARS-CoV-2 patients. Anti-S1-RBD IgA and IgG levels are measured in sera collected between day 1 and day 7 after symptoms onset (n=48). Antibody levels are expressed as arbitrary units/ml (AU/ml). Cut-off lines are represented as grey dotted lines.

**Figure S5 (related to Figure 3):**
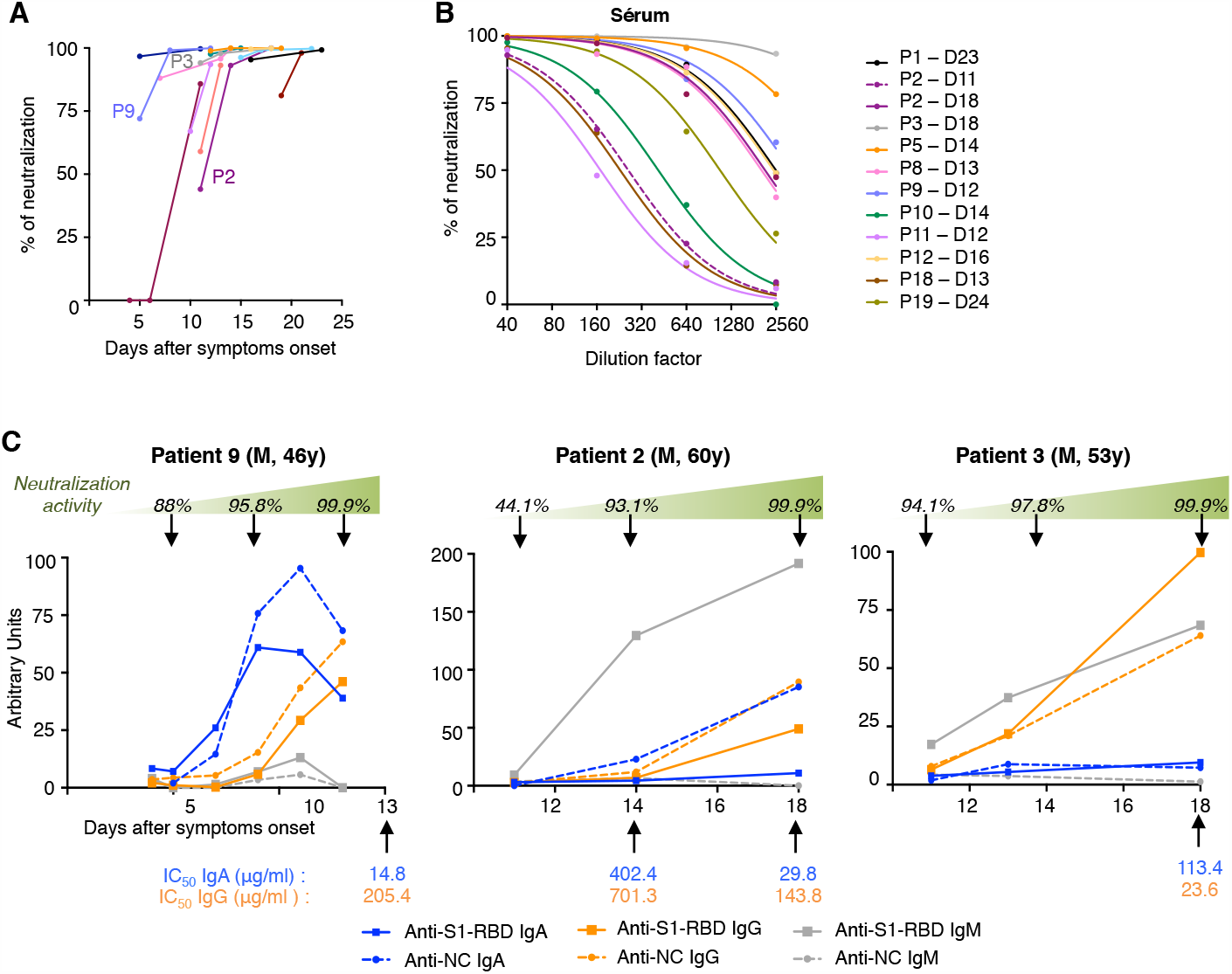
Neutralizing activity of serum from SARS-CoV-2 patients. A. Neutralizing activity of serum (dilution 1:40) measured by pseudovirus neutralization assay in blood collected at indicated time points after symptoms onset in 12 SARS-CoV-2 infected patients. Each colored line represents one patient. Color code is with panel B. B. Neutralizing activity of 12 sera measured by pseudovirus neutralization assay at different indicated of serum. Samples used for this analysis were collected between day 11 and 24 after symptoms onset. C. Three representative immunological evolution profiles. Longitudinal evolution of specific antibody levels (curves) and of neutralizing activity in serum measured at indicated time points (top arrows) are compared. IC_50_ values measured in IgA and IgG purified at indicated time points are also indicated (bottom arrows).

**Table S1:**
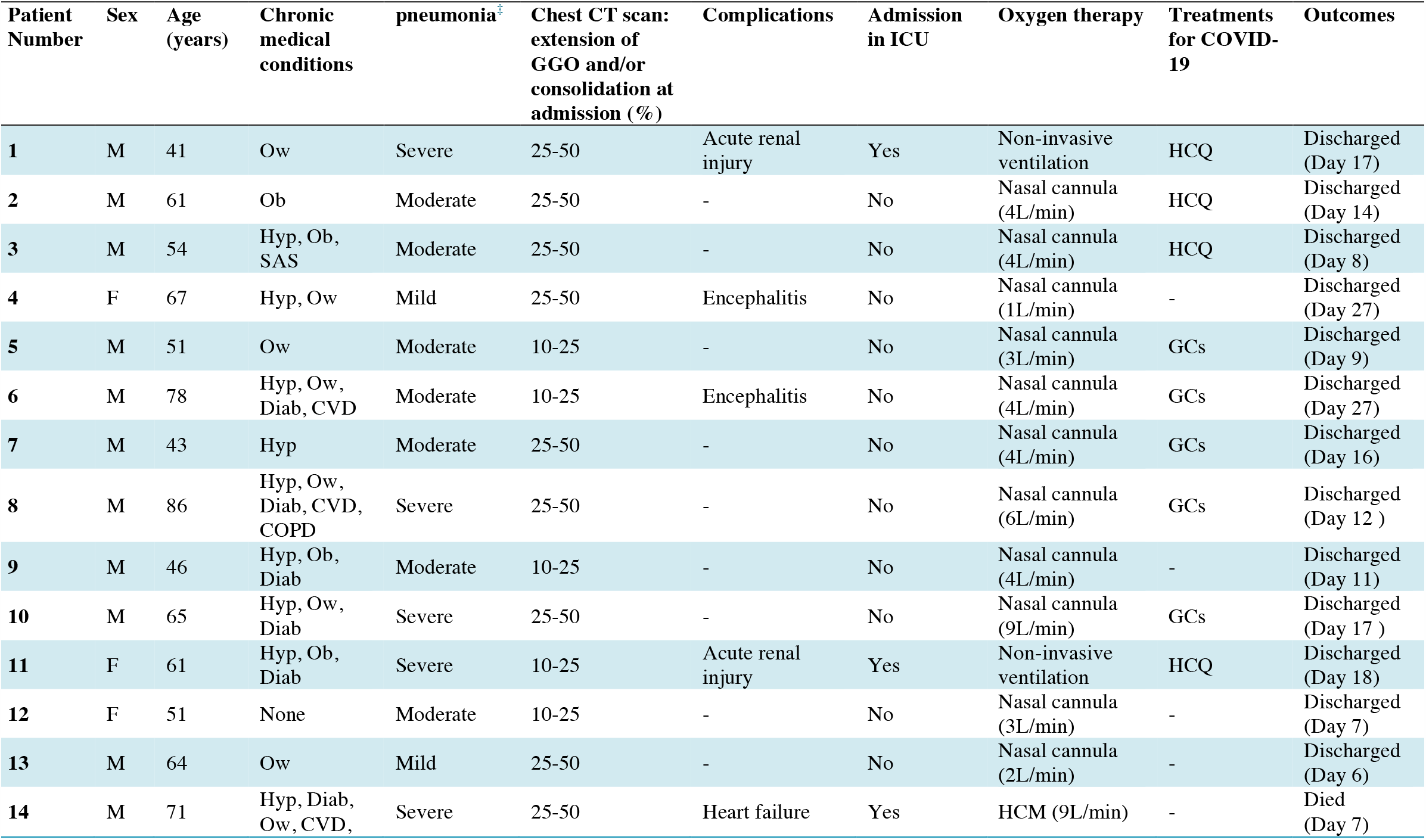
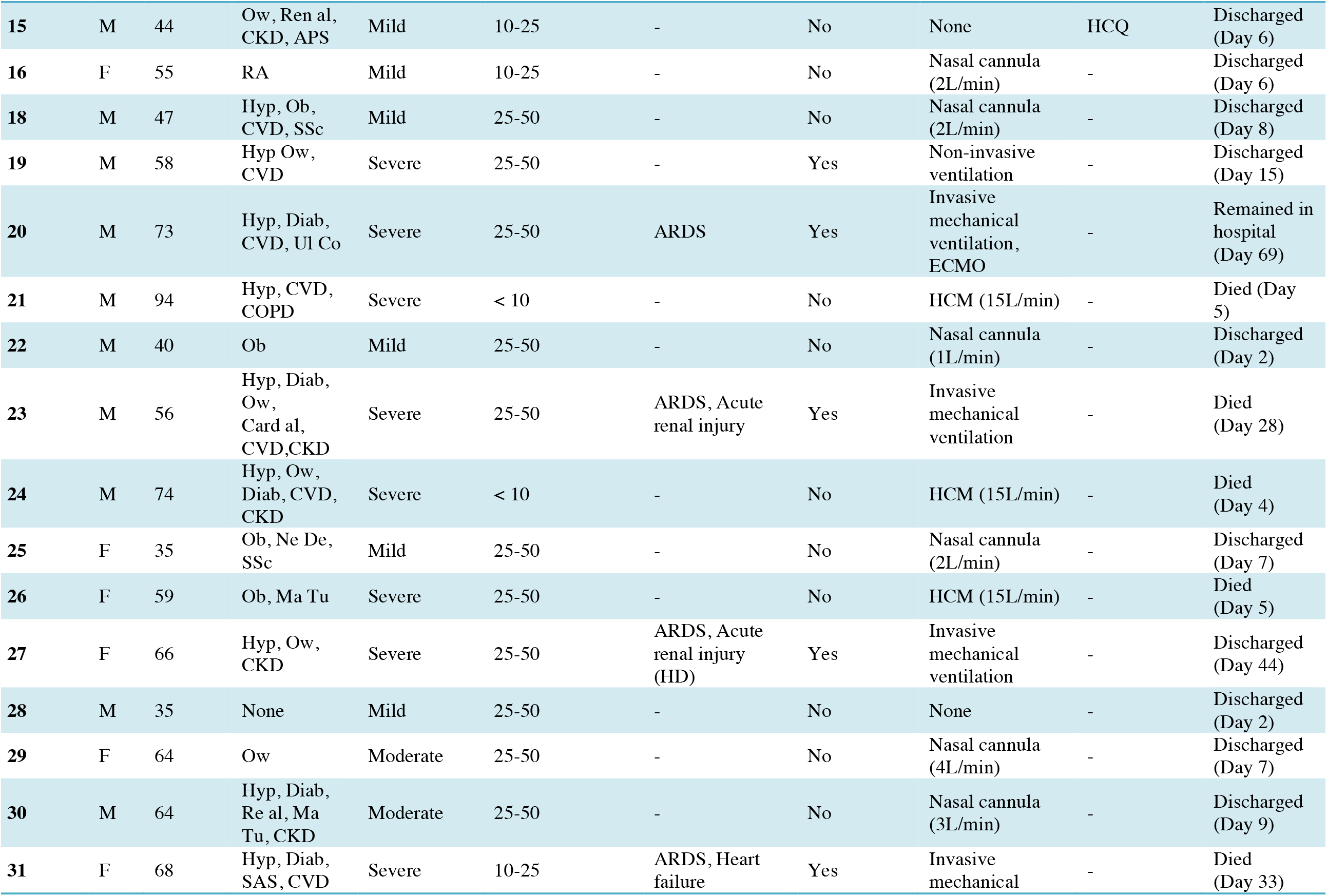
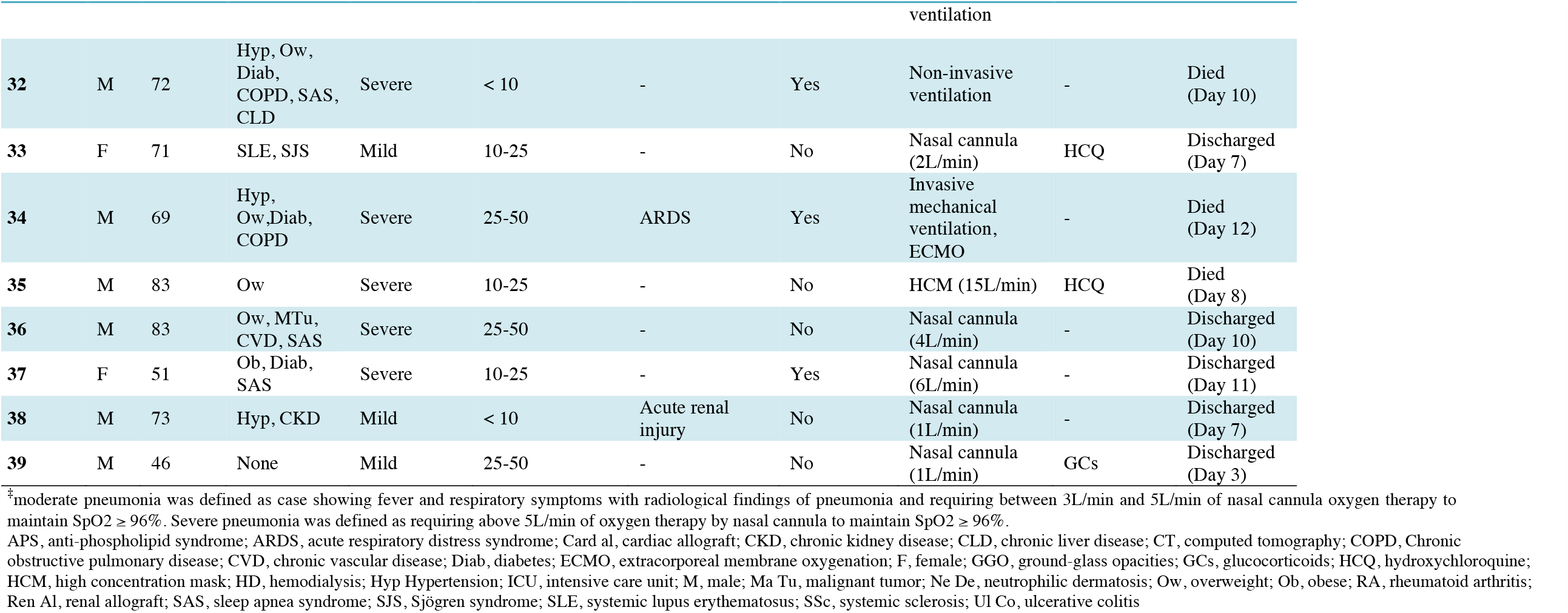
Demographics, baseline characteristics, treatment and outcome of 38 patients with COVID-19 assessed for blood plasmablasts.

**Table S2:** Demographics and baseline characteristics of patients with COVID-19.

|  | **Patients**<br>**(N=135)** |
| --- | --- |
| Women | 55 (41) |
| Age, years, median (quartiles) | 61.3 (49.7 – 72.0) |
| **Chronic medical illness** |  |
| Coronary heart disease or cardiovascular disease | 37 (27) |
| Diabetes | 43 (32) |
| *Body mass index (kg/m<sup>2</sup>)* |  |
| underweight ($\leq 18.5$) | 3 (2) |
| normal (18.5-25) | 43 (32) |
| overweight (25-30) | 48 (36) |
| obesity ($\geq 30$) | 41 (30) |
| Hypertension | 64 (47) |
| Immunocompromised* | 28 (21) |
| Malignant tumor | 12 (9) |
| Nervous system disease | 15 (11) |
| Chronic obstructive lung disease | 12 (9) |
| Chronic kidney disease | 22 (16) |
| Chronic liver disease | 10 (7) |
| *smoking habits* |  |
| never smoker | 78 (58) |
| ex-smoker | 52 (38) |
| daily-smoker | 5 (4) |
| Past history of thrombosis | 14 (10) |
| arterial thrombosis | 5 (4) |
| venous thrombosis | 10 (7) |
| **Treatment regimen** |  |
| Long-term prednisone use | 22 (16) |
| Long-term prednisone $\geq 10$ mg/j | 10 (7) |
| Long-term Immunosuppressive agent use<sup>†</sup> | 15 (11) |
| Nonsteroidal anti-inflammatory drugs | 0 (0) |
| Recent chemotherapy for cancer | 7 (5) |
| angiotensin converting enzyme inhibitor | 28 (21) |
| angiotensin II receptor blockers | 20 (15) |
| Oral anticoagulant | 18 (13) |

Values are expressed as n (%), unless stated otherwise.
\*including cardiac, liver or kidney allograft (n=9), hematopoietic stem cell transplantation (n=1), chemotherapy for cancer (n=6) or immunosuppressant for auto-immune disease (n=12)
<sup>†</sup>Excluding antimalarials and prednisone. Immunosuppressant therapy was mycophenolate mofetil for 9 patients, tacrolimus or cyclosporine for 9, methotrexate for 3, everolimus for 2, rituximab for one and ustekinumab for one.

**Table S3:**
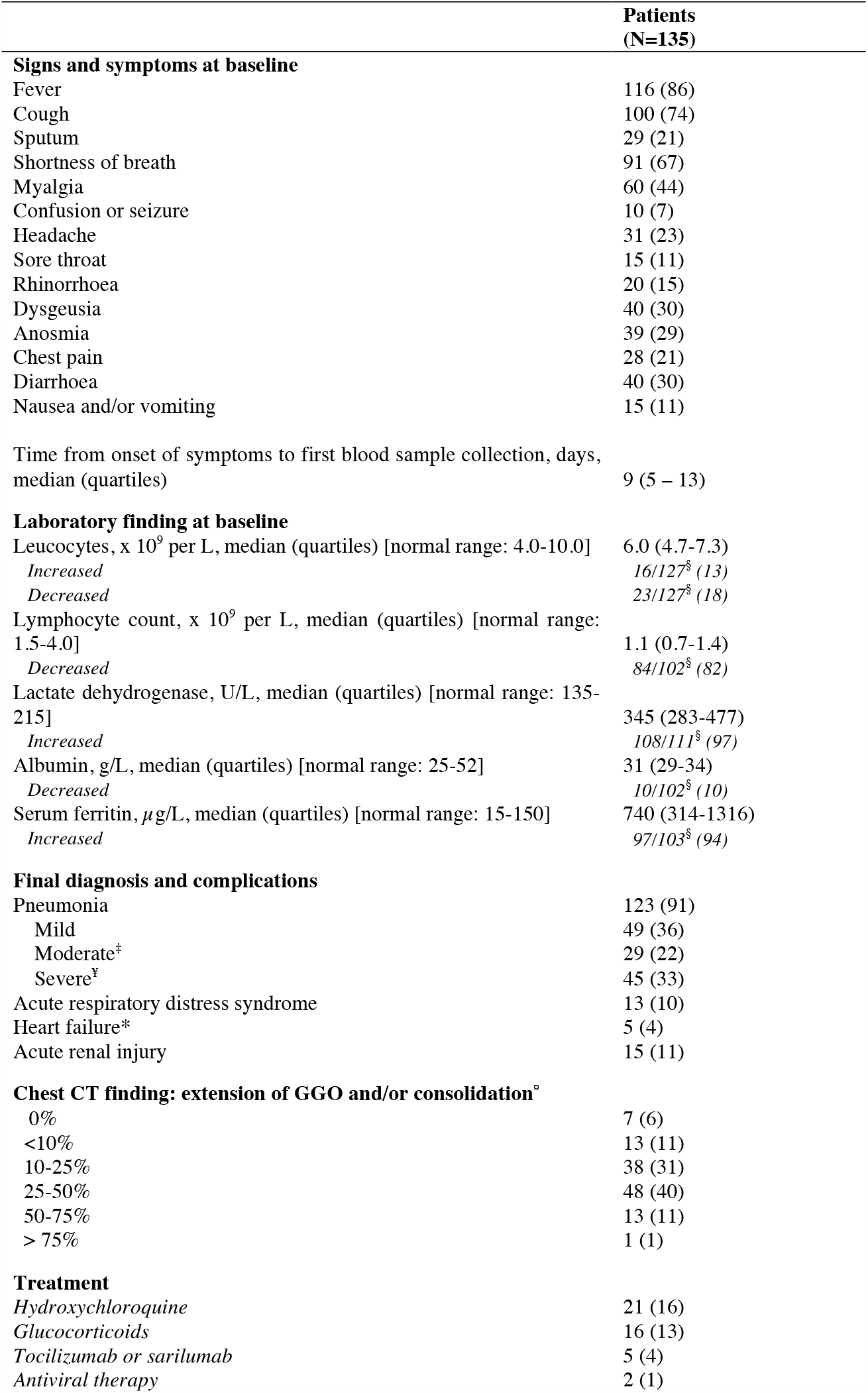
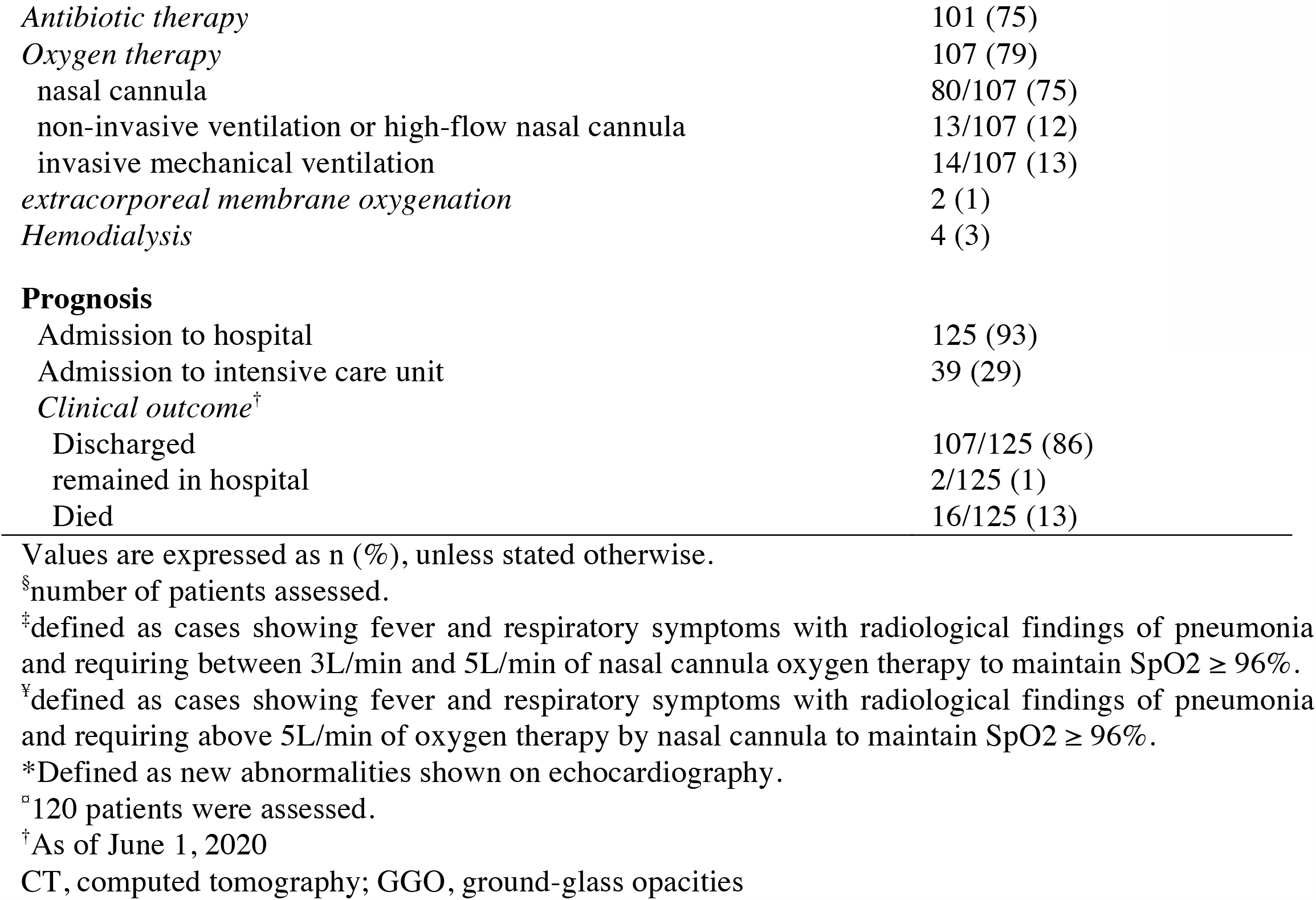
Clinical characteristics, laboratory results, treatment and outcome of patients with COVID-19.

**Table S4:**
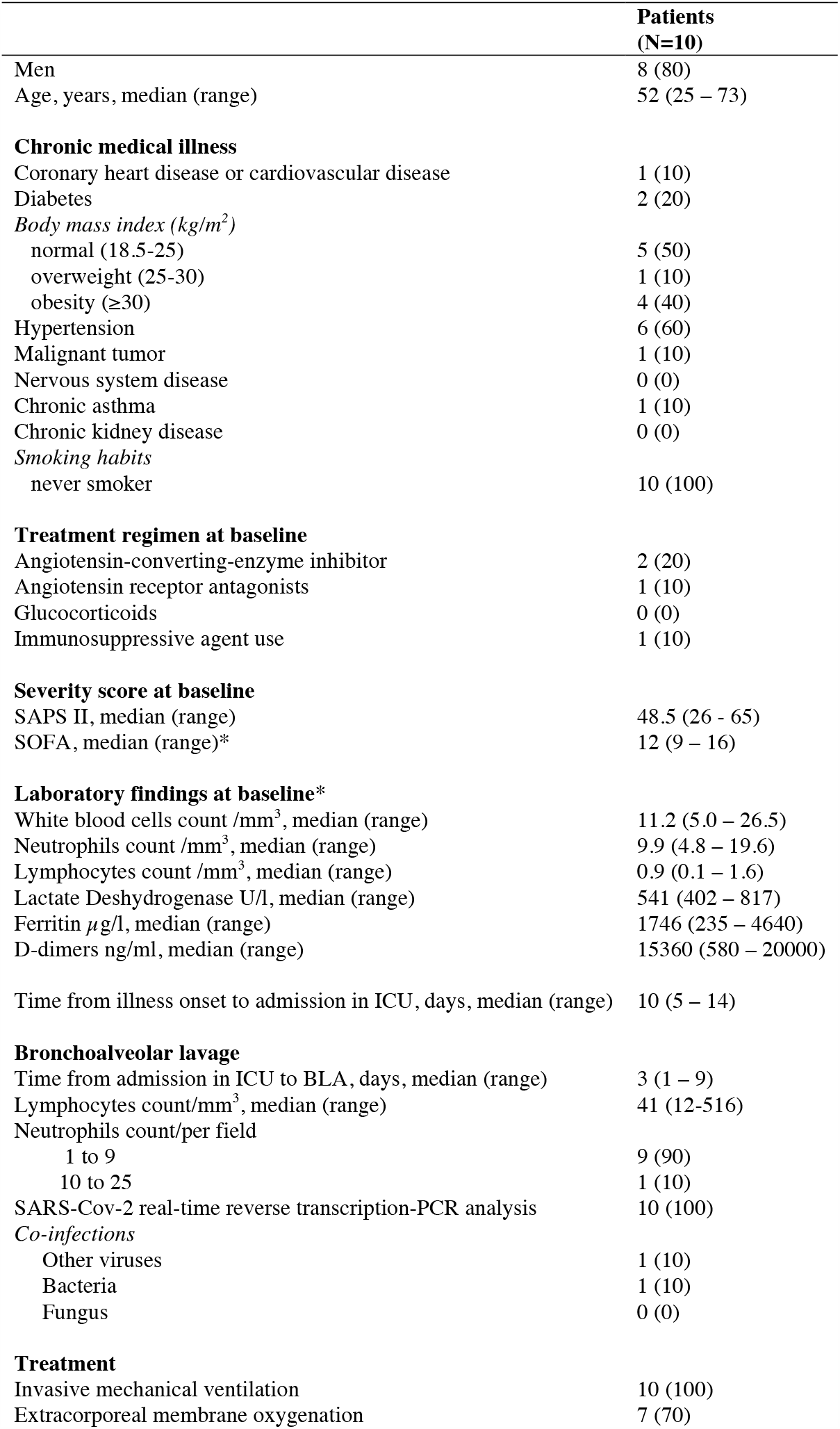
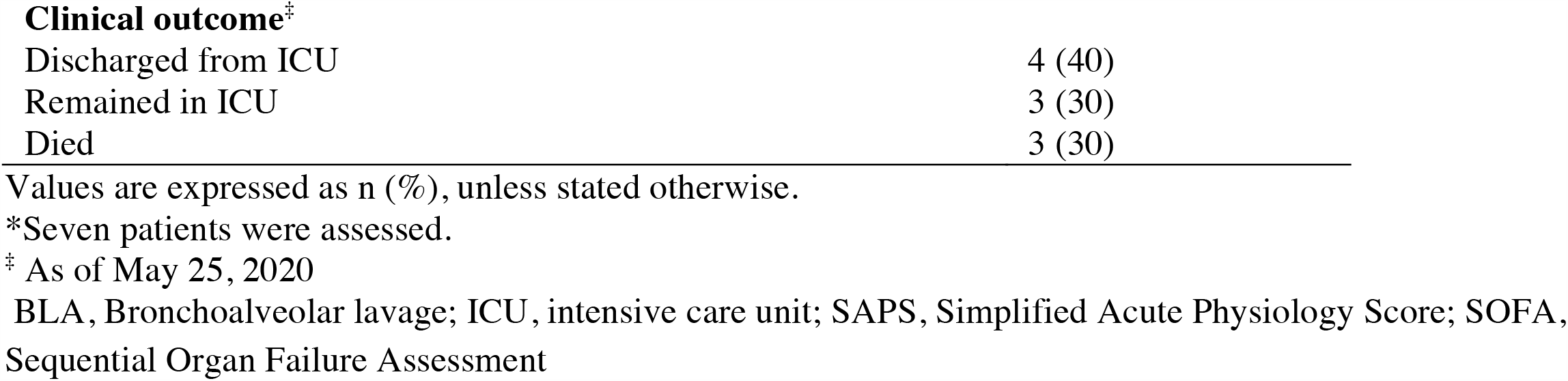
Demographics, baseline characteristics, treatment and outcome of patients with acute respiratory distress syndrome during the course of COVID-19.

|  | **Patients**<br>**(N=10)** |
| --- | --- |
| Men | 8 (80) |
| Age, years, median (range) | 52 (25 – 73) |
| **Chronic medical illness** |  |
| Coronary heart disease or cardiovascular disease | 1 (10) |
| Diabetes | 2 (20) |
| *Body mass index (kg/m<sup>2</sup>)* |  |
| normal (18.5-25) | 5 (50) |
| overweight (25-30) | 1 (10) |
| obesity (≥30) | 4 (40) |
| Hypertension | 6 (60) |
| Malignant tumor | 1 (10) |
| Nervous system disease | 0 (0) |
| Chronic asthma | 1 (10) |
| Chronic kidney disease | 0 (0) |
| *Smoking habits* |  |
| never smoker | 10 (100) |
| **Treatment regimen at baseline** |  |
| Angiotensin-converting-enzyme inhibitor | 2 (20) |
| Angiotensin receptor antagonists | 1 (10) |
| Glucocorticoids | 0 (0) |
| Immunosuppressive agent use | 1 (10) |
| **Severity score at baseline** |  |
| SAPS II, median (range) | 48.5 (26 – 65) |
| SOFA, median (range)* | 12 (9 – 16) |
| **Laboratory findings at baseline\*** |  |
| White blood cells count /mm<sup>3</sup>, median (range) | 11.2 (5.0 – 26.5) |
| Neutrophils count /mm<sup>3</sup>, median (range) | 9.9 (4.8 – 19.6) |
| Lymphocytes count /mm<sup>3</sup>, median (range) | 0.9 (0.1 – 1.6) |
| Lactate Deshydrogenase U/l, median (range) | 541 (402 – 817) |
| Ferritin μg/l, median (range) | 1746 (235 – 4640) |
| D-dimers ng/ml, median (range) | 15360 (580 – 20000) |
| Time from illness onset to admission in ICU, days, median (range) | 10 (5 – 14) |
| **Bronchoalveolar lavage** |  |
| Time from admission in ICU to BLA, days, median (range) | 3 (1 – 9) |
| Lymphocytes count/mm<sup>3</sup>, median (range) | 41 (12-516) |
| Neutrophils count/per field |  |
| 1 to 9 | 9 (90) |
| 10 to 25 | 1 (10) |
| SARS-Cov-2 real-time reverse transcription-PCR analysis | 10 (100) |
| *Co-infections* |  |
| Other viruses | 1 (10) |
| Bacteria | 1 (10) |
| Fungus | 0 (0) |
| **Treatment** |  |
| Invasive mechanical ventilation | 10 (100) |
| Extracorporeal membrane oxygenation | 7 (70) |
| **Clinical outcome<sup>‡</sup>** |  |
| Discharged from ICU | 4 (40) |
| Remained in ICU | 3 (30) |
| Died | 3 (30) |

Values are expressed as n (%), unless stated otherwise.
\*Seven patients were assessed.
<sup>‡</sup> As of May 25, 2020
BLA, Bronchoalveolar lavage; ICU, intensive care unit; SAPS, Simplified Acute Physiology Score; SOFA, Sequential Organ Failure Assessment

**Table S5:** Human Antibodies used for B and T cell phenotyping.

|  |  |
| --- | --- |
| Anti-hCD3 APC-H7 (clone SK7) | BD Biosciences |
| Anti-hCD3 A700 (clone UCHT1) | BD Biosciences |
| Anti-hCD4 PerCP-Cy5.5 (clone SK3) | BD Biosciences |
| Anti-hCD10 APC (clone HI10a) | BD Biosciences |
| Anti-hCD14 A700 (clone M5E2) | BD Biosciences |
| Anti-hCD19 FITC (clone 4G7) | BD Biosciences |
| Anti-hCD19 Bv510 (clone SJ25C1) | BD Biosciences |
| Anti-hCD19 eF450 (clone HIB19) | eBioscience |
| Anti-hCD21 PE (clone B-ly4) | BD Biosciences |
| Anti-hCD27 PeCy7 (clone M-T271) | BD Biosciences |
| Anti-CD38 PerCPCy5.5 (clone HIT2) | BD Biosciences |
| Anti-hCD38 bv510 (clone HIT2) | BD Biosciences |
| Anti-hCCR10 PE (clone 314305) | R&D Systems |
| Anti-hCXCR5 bv421 (clone RF8B2) | BD Biosciences |
| Anti-hKi67 PerCP-Cy5.5 (clone B56) | BD Biosciences |
| Anti-hIgA FITC | Jackson Labs |
| Anti-hIgA PE | Jackson Labs |
| Anti-IgA1 FITC | Southern Biotech |
| Anti-IgA2 A647 | Southern Biotech |
| Anti-hIgD bv421 (clone IA6-2) | BD Biosciences |
| Anti-hIgG A647 | Jackson Labs |
| Anti-hIgM PerCP5.5 | BD Biosciences |
| Anti-hPD-1 bv510 (clone EH12.1) | BD Biosciences |

